# For-Profit Growth and Academic Decline: A Retrospective Nationwide Assessment of Brazilian Medical Schools

**DOI:** 10.1101/2025.05.02.25326906

**Authors:** Bruno B. Andrade, Klauss Villalva-Serra, Rodrigo C. Menezes, Luiz F. Quintanilha, Katia M. Avena

**Affiliations:** Instituto MONSTER de Ensino, Assistência, Pesquisa e Desenvolvimento Tecnológico em Saúde, Salvador, Brazil; Laboratório de Pesquisa Clínica e Translacional, Instituto Gonçalo Moniz, Fundação Oswaldo Cruz, Salvador, Brazil

**Keywords:** Medical Education, Brazil, Academic Performance, Education Business, Undergraduate training

## Abstract

**Background:** The rapid and predominantly for-profit expansion of medical schools in Brazil over the past decade has raised widespread concerns about the erosion of academic standards in medical education.

**Methods:** This nationwide, retrospective study analyzed academic performance indicators from all Brazilian medical schools participating in the 2013, 2016, 2019, and 2023 cycles of the Exame Nacional de Desempenho dos Estudantes (ENADE)—a standardized national exam used to assess students’ knowledge at the end of undergraduate programs. We also included the Indicador de Diferença entre os Desempenhos Observado e Esperado (IDD), which estimates the value added by institutions by comparing student performance at graduation with their academic background at entry. Data were sourced from publicly available datasets provided by the Brazilian Ministry of Education. We compared trends across public, non-profit private, and for-profit private institutions, using descriptive statistics, non-parametric tests, correlation analysis, and Bayesian mixed-effects regression models to assess the impact of institutional category and class size on academic performance.

**Results:** The number of for-profit medical schools in Brazil nearly tripled between 2013 and 2023. These institutions consistently demonstrated lower ENADE scores compared to public and non-profit peers. Although IDD scores showed some early gains, they declined significantly in 2023, particularly among new medical schools taking the ENADE for the first time—most of which were for-profit. Larger class sizes were negatively correlated with both ENADE and IDD scores.

Regression models showed that public institutions outperformed for-profit schools by an average margin of more than 21 ENADE points, while class size emerged as a modest but statistically significant negative predictor of IDD.

**Conclusion:** Our findings reveal that the unregulated expansion of for-profit medical schools in Brazil has been accompanied by a decline in academic performance, as measured by national benchmarks. These patterns suggest a structural misalignment between the commercial logic of expansion and the core educational mission of medical training. Regulatory reforms are urgently needed to realign the growth of medical education with principles of academic quality and social accountability.

## 1 Introduction

Over the past decade, Brazil has experienced the most accelerated expansion of private medical education worldwide, significantly driven by corporate interests, regulatory permissiveness, and the pressures of market dynamics (1–3). This rapid growth, has reshaped the landscape of medical training, exposing critical vulnerabilities - what we previously termed the “dark side of medical education” (2). Mounting concerns have been raised about declining educational quality, inequitable access, and the erosion of academic standards, particularly in for-profit institutions (2,4). However, this phenomenon is not unique to Brazil. Similar trends have emerged across the Global South, especially in Latin American countries such as Ecuador, Peru, and Colombia, and in BRICS countries such as India and South Africa, where systemic challenges are being experienced, including the proliferation of low-quality institutions and the need to address geographic disparities in workforce distribution (2,5,6). These parallels highlight broader systemic issues in global medical education and underscore the relevance of this discussion globally (7). Nevertheless, beyond the proliferation of institutions around the world, a fundamental question remains: what is happening to the academic performance of these students once they graduate?

In Brazil, academic performance in higher education is systematically evaluated through the National Student Performance Exam (*Exame Nacional de Desempenho dos Estudantes* - ENADE), a national examination administered to graduating students across courses, including medicine (8). ENADE assesses the competencies acquired during undergraduate training, functioning as both a national benchmark for program quality and an accountability instrument for educational institutions (9,10). To complement ENADE, the Ministry of Education employs the Indicator of Difference Between Observed and Expected Performance (*Indicador de Diferença entre os Desempenhos Observado e Esperado;* IDD), a measure designed to estimate the educational “value added” by each program by comparing students’ final ENADE scores against their expected performance based on their academic and socioeconomic background at entry (9–11).

Despite growing concerns, the debate surrounding the rapid educational expansion has largely remained speculative, lacking an empirical assessment of its impact on academic outcomes (4–7). Recently, however, the Ministry of Education released the 2023 ENADE and IDD data for health undergraduate courses, covering nearly a thousand classes from public, non-profit, and for-profit medical schools (9). Their report suggests a systemic issue: private, for-profit institutions, particularly those that have scaled aggressively over the past decade, showed consistent patterns of underperformance. These findings are not statistical outliers, but symptoms of a broader educational deterioration affecting thousands of medical students and, by extension, potentially compromising the future of public health in Brazil.

Faced with this evidence, we revisited this issue, emphasizing a “darker side” of medical education. In this follow-up article, we present a decade-long analysis of national performance data to expose the structural decline in academic performance linked to institutional expansion and market-driven medical education. Our analysis begins in 2013, the year in which the Brazilian government launched the *Programa Mais Médicos* (More Doctors Program), a governmental policy designed to address physician shortages and stimulate medical school expansion (12,13). Rather than merely continuing prior discussions, this article aims to quantify and characterize the structural decline in academic outcomes related to the rapid institutional proliferation and the commercialization of medical education. This investigation serves not only as a critical response to an intensifying educational crisis, but also to highlight the urgent need for systemic reforms in Brazil’s medical education landscape.

## 2 Methods

### 2.1 Study Design

This was a retrospective, observational, nationwide study based on secondary data from the Brazilian Ministry of Education. We analyzed institution-level academic performance metrics from four ENADE cycles (2013, 2016, 2019, and 2023) for medical school programs, which are conducted on a tri-annual basis. Our analysis included all cycles performed since the beginning of the *More Doctors* program (12,13).

### 2.2 Data Sources

We used publicly available databases released by the National Institute for Educational Studies and Research Anísio Teixeira (*Instituto Nacional de Estudos e Pesquisas Educacionais Anísio Teixeira -* INEP), the technical arm of Brazil’s Ministry of Education, responsible for overseeing the national evaluation system for higher education. The dataset included institution-level ENADE and IDD scores, administrative category (public, non-profit private, and for-profit private), geographic location, and number of students participating in ENADE per institution. Schools with missing data or classified under non-standard categories, such as binational or in a transitional status, were excluded. All datasets are available on INEP’s website (9).

### 2.3 Outcome Measures

ENADE is a standardized national examination applied to final-year undergraduate students to assess the knowledge and competencies acquired during training in Brazil. Established by Law No. 10,861 in 2004 as part of the Brazilian Higher Education Evaluation System (Sinaes) (8), the exam evaluates students’ performance in relation to the content outlined in the Brazilian National Curricular Guidelines, as well as the development of general and professional competencies and their understanding of contemporary national and global issues. In medical education, ENADE covers areas such as clinical knowledge, public health, ethics, and professional practice (8).

IDD complements ENADE by estimating the “educational value added” of each program. It is calculated by comparing the observed ENADE scores of graduating students to a predicted score generated through regression models that incorporate academic performance at entry, primarily the *Exame Nacional do Ensino Médio* (ENEM), as well as socioeconomic and demographic variables. Both ENADE and IDD are officially reported as continuous scores ranging from 0 to 5 and categorical ordinal classes from 1 (lowest) to 5 (highest), as defined in INEP’s official technical guidelines (8–10).

The assessment follows a triennial cycle, meaning each program is evaluated every three years. Participation in ENADE is mandatory for graduation in the years in which it is applied. Therefore, we used the number of participants in the test as a proxy for total class sizes for each institution. To evaluate whether increased class size was associated with changes in academic performance, we calculated normalized ENADE and IDD scores by dividing the total institutional score by the number of students participating in the examination. For IDD, we used the number of students with both ENADE and Brazilian National High School Exam (Exame Nacional do Ensino Médio - ENEM) data available, as required for its calculation (14). ENEM is a nationwide standardized exam administered by INEP at the end of upper secondary education, and it is widely used as a university entrance examination in Brazil. These normalized metrics were used in correlational and regression analyses to assess the association between institutional scale and student outcomes.

### 2.4 Statistical Analysis

ENADE and IDD scores were linearly transformed to a 0–100 scale to enhance interpretability and facilitate comparison with international benchmarks. Both continuous and grouped versions of the scores were used in the analysis. For categorical analyses, scores were classified into five intervals (20, 40, 60, 80, 100), following the grouping parameters established by the Brazilian Ministry of Education.

Descriptive statistics were calculated for ENADE and IDD scores across administrative categories (public, non-profit private, and for-profit private). The Shapiro–Wilk test was used to assess the normality. As the data were non-normally distributed, differences between groups were evaluated using the non-parametric Kruskal–Wallis test, followed by Dunn’s post hoc test with Bonferroni correction for multiple comparisons. Categorical values were compared using Pearson’s Chi-squared test, while the Spearman test of trend was performed to assess changes in linear trends of university data through this ten-year period. Spearman’s rank correlation coefficient (ρ) was used to assess the monotonic relationships between institutional class size and ENADE scores, class size and IDD scores, and ENADE and IDD scores. Correlation matrices and rank-transformed scatterplots were generated to visualize these associations. All tests were two-sided, and a p-value < 0.05 was considered statistically significant.

To quantify the association between administrative category and performance outcomes, we implemented Bayesian generalized linear mixed-effects regression models using the Integrated Nested Laplace Approximation (INLA) framework (15). Separate models were constructed for ENADE and IDD as dependent variables, with institutional type and log-transformed class size as fixed effects. To account for potential spatial and temporal autocorrelation, we included two random effects: a spatial component modelled using an Intrinsic Conditional Autoregressive (ICAR) structure based on Brazilian states, and a temporal component modelled as a first-order random walk (RW1) over the four ENADE cycles. Posterior means and 95% credible intervals (CrI) were used to interpret the regression coefficients.

Statistical analyses were conducted using RStudio (version 4.3.4).

### 2.5 Ethics Statement

All data used in this study were obtained from publicly available government databases and did not involve identifiable personal information. Therefore, it did not require submission to the Brazilian Research Ethics Committee, as dictated by Resolution No. 510/2016 of the Brazilian Health Council on norms applicable to Human and Social Sciences research.

## 3 Results

The number of institutions participating in the ENADE exam varied across the four evaluation cycles. For this study, we excluded institutions classified under administrative categories outside the scope of public, non-profit private, or for-profit private universities, such as the four “Special” category schools in 2023. Additionally, institutions lacking ENADE and IDD scores were excluded (one in 2013 and four in 2023). After applying these criteria, the final analytical sample comprised 165 institutions in 2013, 176 in 2016, 232 in 2019, and 301 in 2023.

### 3.1 The Expansion Effect: More Schools, More Seats, Lower Scores

Between 2013 and 2023, the number of medical schools in Brazil nearly doubled, rising from 163 to 301 with for-profit private institutions leading this growth. Their representation increased from 30 medical schools (18.4%) in 2013 to 90 (29.8%) in 2023 (p-value for trend = 0.030) (**Table 1**). As shown in **Figure 1A**, this expansion was accompanied by a sharp rise in ENADE participation, especially among for-profit institutions. Simultaneously, ENADE scores stagnated or declined, most notably among for-profit schools (**Figure 1B**). While overall IDD scores showed an upward trajectory from 2013 to 2023, a significant drop was observed in both continuous and class-size-normalized IDD scores between 2019 and 2023 (pairwise p-value < 0.001) (**Table 1**).

**Figure 1.**
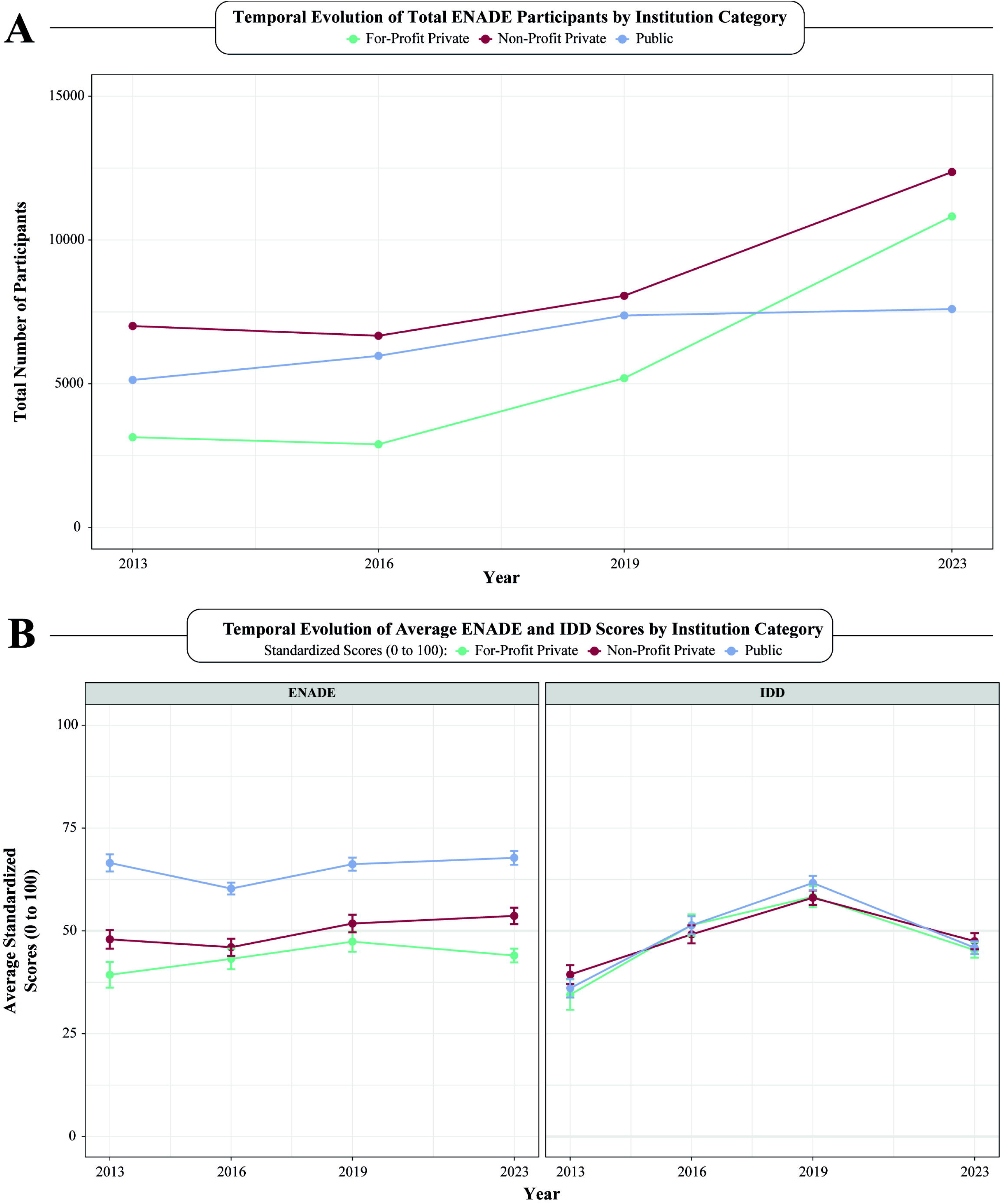
Temporal Evolution of ENADE Participation and Academic Performance by Institution Category. (A) Evolution of total sum of medical students who participated in the ENADE exam, according to institution administrative categories, including public (blue), non-profit (red) and for-profit (green) private universities per year (2013 to 2023). (B) Temporal evolution among average ENADE and IDD scores plotted per year (2013 to 2023), according to institution administrative categories. Error bars were used to showcase standard error of ENADE and IDD scores. ENADE: National Student Performance Exam; IDD: Indicator of Difference between Observed and Expected Academic Performance.

**Table 1.** Brazilian Medical School’s Standardized Scores and Participants from 2013-2023.

| Variables | 2013<br>(n=163) | 2016<br>(n=176) | 2019<br>(n=232) | 2023<br>(n=301) | Global<br>p-value | Trend<br>p-value | 2013 vs 2016<br>p-value | 2016 vs<br>2019<br>p-value | 2019 vs 2023<br>p-value |
| --- | --- | --- | --- | --- | --- | --- | --- | --- | --- |
| **Institution Category** |  |  |  |  | **0.030** | **0.031** | 0.804 | 0.492 | 0.326 |
| For-Profit Private | 30 (18.4%) | 32 (18.2%) | 53 (22.8%) | 90 (29.9%) |  |  |  |  |  |
| Non-Profit Private | 69 (42.3%) | 69 (39.2%) | 79 (34.1%) | 99 (32.9%) |  |  |  |  |  |
| Public | 64 (39.3%) | 75 (42.6%) | 100 (43.1%) | 112 (37.2%) |  |  |  |  |  |
| **ENADE Metric** |  |  |  |  |  |  |  |  |  |
| Participants (Class Size) | 88.0 [60.5;110] | 83.5 [54.8;106] | 85.0 [53.8;113] | 90.0 [53.0;131] | 0.210 | 0.187 | 0.535 | 0.674 | 0.289 |
| Continuous Score | 55.7[40.4;69.0] | 53.6 [41.9;64.2] | 59.3 [44.6;71.7] | 56.5 [42.9;71.2] | **0.011** | **0.038** | 0.354 | **0.005** | 0.515 |
| Class-Size Normalized Score | 0.61 [0.36;1.02] | 0.64 [0.41;0.94] | 0.67 [0.43;1.07] | 0.61 [0.35;1.13] | 0.358 | 0.858 | 0.861 | 0.458 | 0.402 |
| **IDD Metric** |  |  |  |  |  |  |  |  |  |
| Participants | 86.0 [57.5;120] | 72.0 [48.0;94.2] | 81.0 [49.0;108] | 85.0 [49.0;123] | **0.002** | 0.451 | **0.003** | 0.104 | 0.122 |
| Continuous Score | 37.4 [23.7;49.5] | 52.5 [42.1;61.1] | 62.6 [53.8;69.5] | 46.7 [34.7;58.8] | **<0.001** | **0.003** | **<0.001** | **<0.001** | **<0.001** |
| Class-Size Normalized Score | 0.43 [0.21;0.70] | 0.68 [0.46;0.97] | 0.74 [0.50;1.15] | 0.55 [0.31;0.93] | **<0.001** | 0.050 | **<0.001** | 0.126 | **<0.001** |

**Table Note:** This table compares characteristics of Brazilian Medical Schools in a ten-year period from 2013-2023. Data includes standardized score metrics (ENADE and IDD) of Brazilian Medical Schools, as well as test participants and institutional types: public schools, for-profit and non-profit private school. Data is presented as total numbers and frequency (%), as well as Median and interquartile range (IQR). Global and pairwise comparisons being performed with the Kruskal-Wallis test for continuous data and chi-squared test for categorical data. Spearman test for trend was performed to assess changes in trends of university data through this ten-year period.

In the 2023 ENADE cycle, public institutions significantly outperformed both non-profit and for-profit private schools (p-value < 0.001). Non-profit private institutions also achieved higher scores than their for-profit counterparts, reinforcing a persistent and stratified performance gap across institutional categories (**Figure 2** and **Table 2**). In contrast, IDD scores did not significantly differ across the three categories (p-values > 0.70), suggesting that the relative “value added” during medical training, when adjusted for student background, was similar. Nonetheless, the absolute ENADE scores remained much lower for for-profits institutions, highlighting persistent disparities in final academic achievement despite apparent parity in learning gains.

**Figure 2.**
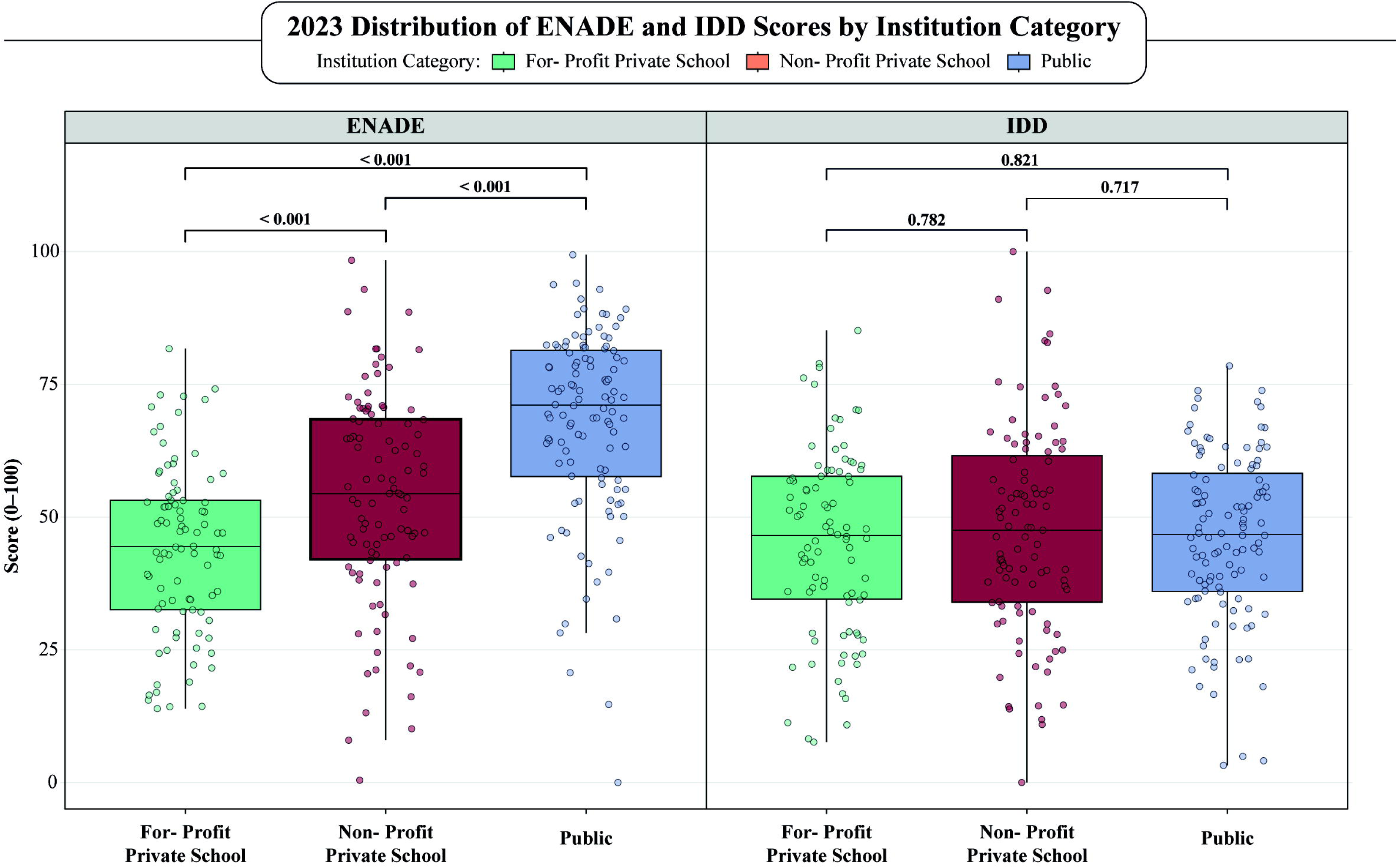
Comparative distribution of ENADE and IDD Scores Across Public, Non-Profit, and For-Profit Medical Schools in 2023. This figure represents box plots showcasing the distribution of ENADE and IDD scores plotted from the last ENADE exam year (2023). Both pairwise and global comparisons were performed, to assess differences in scores between institutional categories, as well as overall differences. Due to non-normal distributions in our data, we used the non-parametric Kruskal-Wallis testing to assess the statistical significance of comparisons. Green represents For-Profit Private Schools; Red represents Non-Profit Private Schools and Blue represents Public Schools. ENADE: National Student Performance Exam; IDD: Indicator of Difference between Observed and Expected Academic Performance.

**Table 2.** Standardized Scores and Participants of Brazilian Medical Schools According to Administrative Category.

| Variables | For-Profit Private<br>(n=90) | Non-Profit Private<br>(n=99) | Public<br>(n=112) | Global<br>p-value | Pairwise Comparisons |  |  |
| --- | --- | --- | --- | --- | --- | --- | --- |
|  |  |  |  |  | For-Profit vs<br>Non-Profit<br>Private Schools | For-Profit Private vs<br>Public Schools | Non-Profit Private vs<br>Public Schools |
| Enade Metric |  |  |  |  |  |  |  |
| Participants (Class Size) | 110 [66.2;156] | 112 [79.5;164] | 56.5 [36.8;86.0] | <0.001 | 0.498 | <0.001 | <0.001 |
| Continous Score | 44.4 [32.5;53.2] | 54.4 [42.1;68.4] | 71.1 [57.6;81.4] | <0.001 | <0.001 | <0.001 | <0.001 |
| Normalized Score | 0.39 [0.25;0.62] | 0.47 [0.31;0.71] | 1.24 [0.80;1.89] | <0.001 | 0.145 | <0.001 | <0.001 |
| IDD Metric |  |  |  |  |  |  |  |
| Participants | 102 [56.0;135] | 106 [78.0;152] | 54.5 [33.8;84.2] | <0.001 | 0.283 | <0.001 | <0.001 |
| Continous Score | 46.5 [34.6;57.7] | 47.5 [34.0;61.6] | 46.7 [36.0;58.2] | 0.836 | 0.782 | 0.782 | 0.782 |
| Class-Size Normalized Score | 0.47 [0.22;0.82] | 0.42 [0.27;0.63] | 0.89 [0.52;1.33] | <0.001 | 0.387 | <0.001 | <0.001 |

**Table Note:** This table compares standardized score metrics (ENADE and IDD) of Brazilian Medical Schools according to their institutional types: public schools, for- profit and non-profit private school. Data is presented as Median and interquartile range (IQR), with global and pairwise comparisons being performed with the Kruskal- Wallis test.

### 3.2 Comparison of Class-Size Normalized Standardized Testing Scores

Given that both ENADE and IDD scores are calculated as raw institutional averages based on the performance of all participating students, we adjusted these metrics by normalizing each institution’s score according to its estimated class size. This adjustment allowed us to examine whether graduating cohort size alters the relationship between an institution’s administrative category and its ENADE and IDD performance.

Using data from the 2023 evaluation cycle, the class-size normalization revealed a pronounced performance advantage for public universities. Their median class-size-adjusted ENADE score was more than twice as high as that of either private group, with statistically significant differences observed when compared to both for-profit (p < 0.001) and non-profit (p < 0.001) institutions (Table 2 and Figure 3). In contrast, no significant difference was found between the two private categories (p = 0.145), indicating that profit status alone does not substantially affect per-student ENADE performance once cohort size is accounted for. A similar trend was observed for IDD scores: public institutions again achieved significantly higher normalized values than both for-profit and non-profit schools (p < 0.001 for both comparisons), while no significant difference emerged between the private categories (p = 0.853).

**Figure 3.**
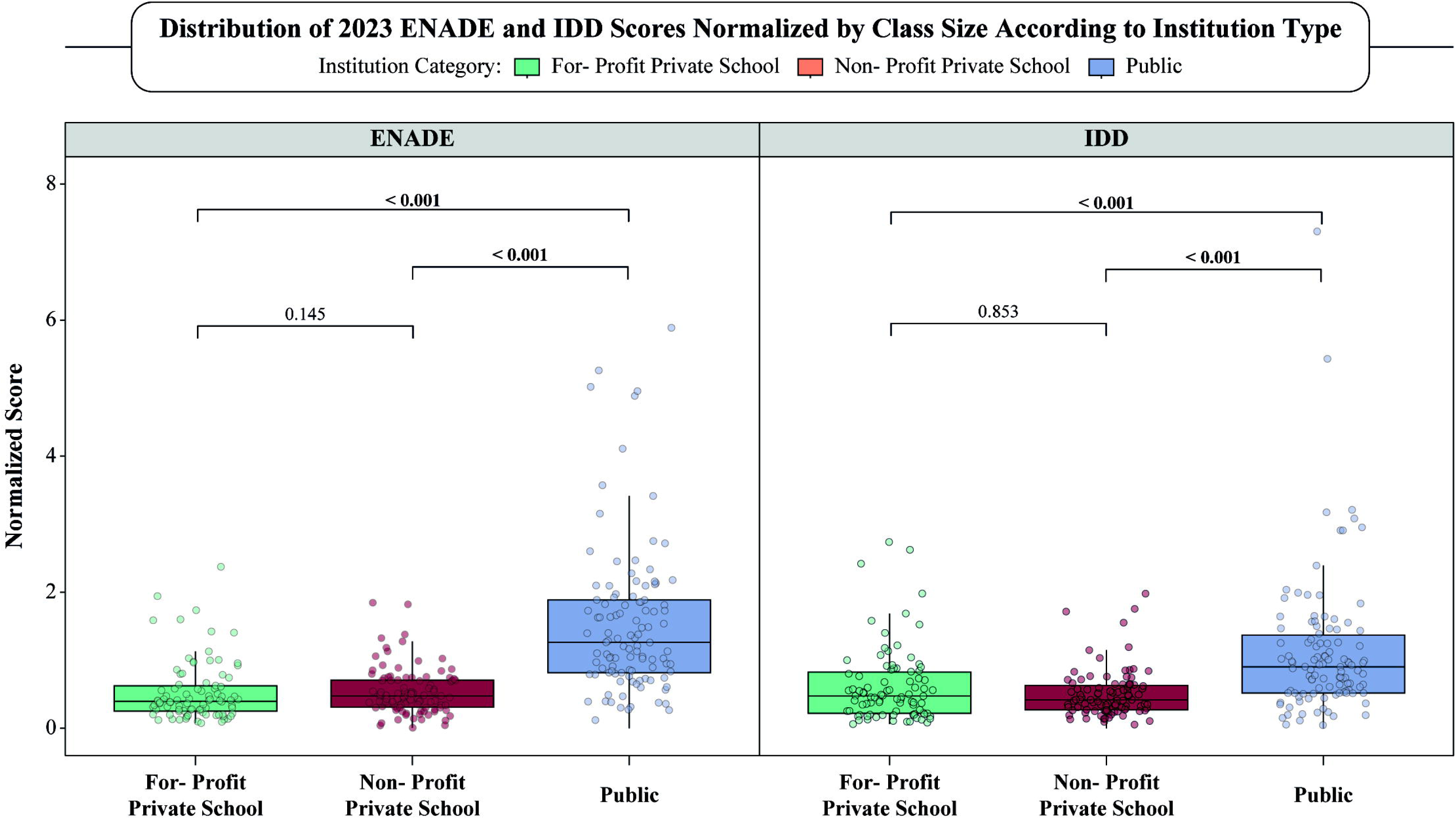
Class-Size-Normalized ENADE and IDD Scores (2023) Across Brazilian Institution Type. Box-and-whisker plots depict the distribution of ENADE (left panel) and IDD (right panel) scores for the 2023 examination year after normalizing each institution’s raw score by the number of students who sat the test (score ÷ participants). Points represent individual universities; boxes show the median and inter-quartile range (IQR) are represented as whiskers. Pairwise comparisons between institution categories were evaluated with the Kruskal–Wallis test followed by Bonferroni-adjustments, owing to non-normal score distributions; corresponding p-values are annotated above the brackets. Colors identify institution type: green = for-profit private, orange = non-profit private, blue = public.

### 3.3 Shifts in Score Distribution and Decline of Educational Value Added

Figure 4A shows the evolution of ENADE and IDD scores across four evaluation cycles. Considering only the most recent tests, from 2019 to 2023, the proportion of institutions in the lower performance brackets (20th and 40th percentiles) increased from 14% to 20%, while the share in the higher-performing bands (80th and 100th percentiles) declined from 51% to 46%. However, while these changes suggest a redistribution, the variations over the period evaluated do not clearly show a trend in the evolution of scores. With the IDD, a similar scenario arose. The proportion of institutions in the top two quintiles fell sharply from 60% to 25%, while those in the lowest two quintiles surged from 10% to 33%.

**Figure 4.**
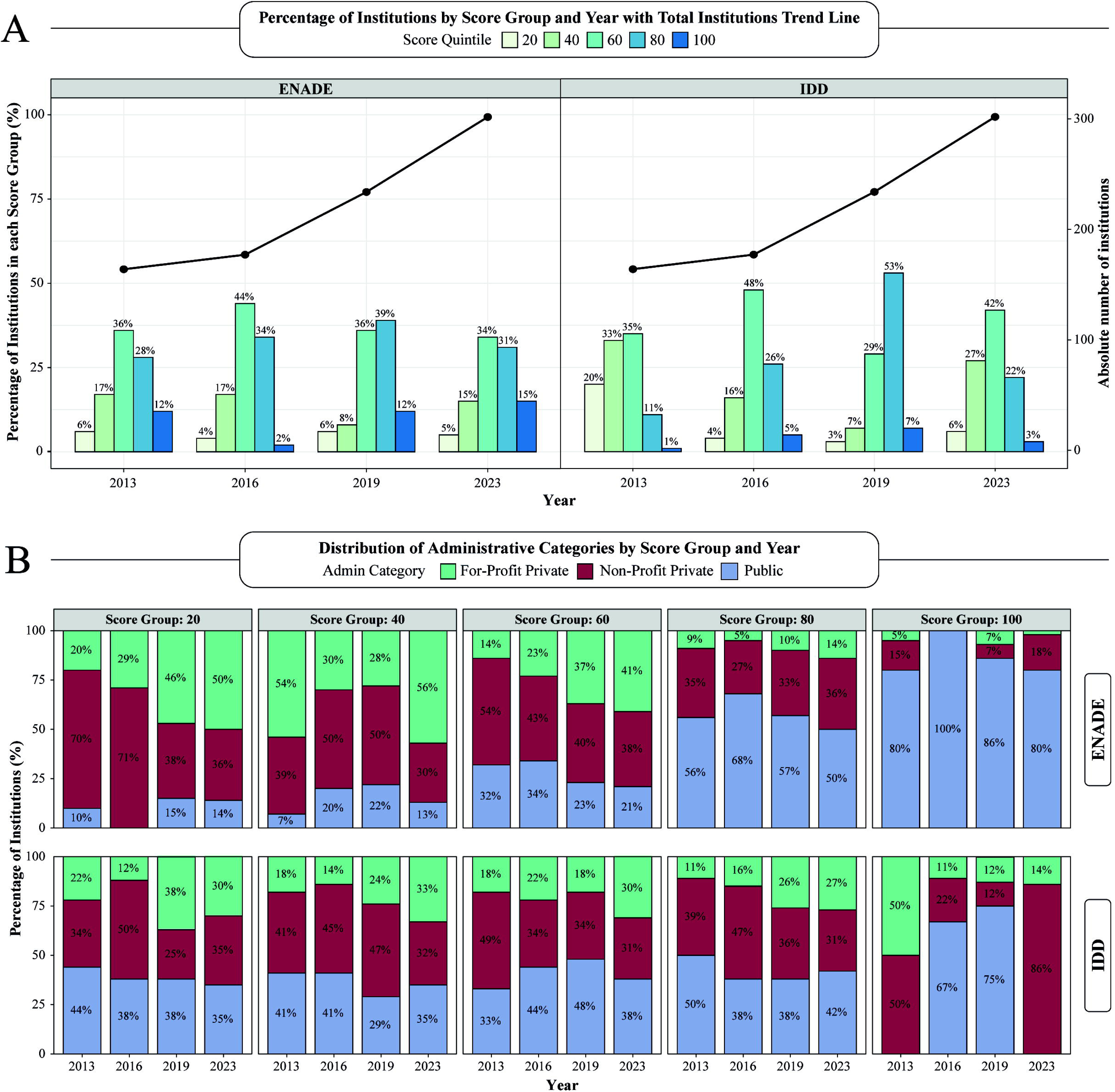
Distribution of Medical Schools by ENADE and IDD Score Group and Year (2013– 2023). **A)** This figure illustrates the longitudinal distribution of institutions by ENADE and IDD score group, 20^th^, 40^th^, 60^th^, 80^th^, and 100^th^, across four ENADE exam years (2013, 2016, 2019, and 2023). Bars denote the percentage of institutions falling into each specific score group (left Y-axis), while the superimposed black line plots the overall count of institutions in each year (right Y-axis), highlighting a substantial growth in the total number of participating schools over time. B) Stacked Bar Chart illustrating the distribution of ENADE and IDD score groups across categories of institutional administration over a ten-year period, including 4 standardized testing cycles. ENADE: National Student Performance Exam; IDD: Indicator of Difference between Observed and Expected Academic Performance.

Furthermore, we analyzed how ENADE and IDD score groups are distributed across institutional types. Figure 4B shows that for-profit universities increasingly concentrated in the lower ENADE score quintiles, representing more than half of the institutions in the 20th and 40th percentiles by 2023, and 41% in the 60th percentile. Their share of the bottom quintile rose from 20% in 2013 to 50% in 2023, while non-profit private institutions declined from 70% to 36%, and public institutions consistently remained below 15%. At the other extreme, public universities overwhelmingly occupy the top-value ENADE bands, contributing 80-100% of institutions in the 100th percentiles over the entire decade, and consistently more than 50% of those in the 80th percentile.

Similar patterns were found when assessing IDD performance public institutions accounted for 44% of the lowest-scoring quintile in 2013, a share that fell to 35% by 2023, while for-profit institutions increased from 22% to 30%, and non-profit schools remained relatively stable around one-third. However, even though public institutions held a relatively stable rate among the 80^th^ tier, representing around 40-50%, their presence in the top-value quintile collapsed, from 67 to 75% in 2016-2019 to 0% in 2023, leaving non-profit private institutions as the majority of highest IDD scorers (Figure 3B).

### 3.4 ENADE and IDD Performance: First-Time vs. Returning Institutions

To examine whether prior ENADE exposure shaped performance trajectories, we divided medical school campuses into two cohorts: first-time participants, which inaugural ENADE cycle occurred in one of the cycle years (2013, 2016, 2019, or 2023), and returning participants, with at least one prior ENADE appearance. This categorization allowed us to compare “debut” outcomes with the longitudinal stability (or drift) of established institutions.

Among first-time participants, both ENADE and IDD quintile distributions changed significantly across cycles (global p-value < 0.001), but only IDD exhibited a sustained linear trend (p-value = 0.003). The only significant pairwise shift for both metrics occurred between 2019 and 2023 (p-value = 0.001), when the share in the lowest quintiles (20th + 40th) surged; ENADE from 5.3% to 33.8%, IDD from 8.7% to 38.8%, while the top-value quintiles (80th + 100th) subsequently declined (**Table 3**).

**Table 3.** Score-Group Distribution of First-Time ENADE Participants by Exam Year.

| Score Groups | 2013<br>(n=163) | 2016<br>(n=15) | 2019<br>(n=57) | 2023<br>(n=80) | Global<br>p-value | Trend<br>p-value | 2013 vs 2016<br>p-value | 2016 vs 2019<br>p-value | 2019 vs 2023<br>p-value |
| --- | --- | --- | --- | --- | --- | --- | --- | --- | --- |
| **ENADE** |  |  |  |  | **<0.001** | 0.263 | 0.906 | 0.183 | **0.001** |
| 20th | 10 (6.1 %) | 0 (0.0 %) | 2 (3.5 %) | 8 (10.0 %) |  |  |  |  |  |
| 40th | 28 (17.2 %) | 3 (20.0 %) | 1 (1.8 %) | 19 (23.8 %) |  |  |  |  |  |
| 60th | 59 (36.2 %) | 7 (46.7 %) | 21 (36.8 %) | 31 (38.8 %) |  |  |  |  |  |
| 80th | 46 (28.2 %) | 4 (26.7 %) | 24 (42.1 %) | 16 (20.0 %) |  |  |  |  |  |
| 100th | 20 (12.3 %) | 1 (6.7 %) | 9 (15.8 %) | 6 (7.5 %) |  |  |  |  |  |
| **IDD** |  |  |  |  | **<0.001** | **0.003** | **0.017** | **0.048** | **<0.001** |
| 20th | 32 (19.6 %) | 0 (0.0 %) | 1 (1.8 %) | 8 (10.0 %) |  |  |  |  |  |
| 40th | 54 (33.1 %) | 3 (20.0 %) | 4 (7.0 %) | 23 (28.8 %) |  |  |  |  |  |
| 60th | 57 (35.0 %) | 6 (40.0 %) | 8 (14.0 %) | 32 (40.0 %) |  |  |  |  |  |
| 80th | 18 (11.0 %) | 4 (26.7 %) | 36 (63.2 %) | 15 (18.8 %) |  |  |  |  |  |
| 100th | 2 (1.2 %) | 2 (13.3 %) | 8 (14.0 %) | 2 (2.5 %) |  |  |  |  |  |

**Table Note:** Counts and percentages of campuses making their inaugural ENADE appearance in each focal cycle (2013, 2016, 2019, 2023), stratified by ENADE and IDD quintiles. Global p-values ( $\chi^2$ ) test differences across all four years; trend p-values derive from the Cochran–Armitage test for ordered change; pairwise p-values compare adjacent cycles (2013 vs 16, 2016 vs 19, 2019 vs 23); ENADE: National Student Performance Exam; IDD: Indicator of Difference between Observed and Expected Academic Performance.

In returning institutions, ENADE shifts were both globally and linearly significant (p-value < 0.001), with the primary change between 2016 and 2019 (p-value = 0.002) before stabilizing. The top-value ENADE quintile rose steadily from 1.2% to 17.2%, indicating growing performance consolidation among experienced campuses. IDD, by contrast, fluctuated, peaking in 2019 (80th quintile 49.7%) then receding by 2023 (23.1%) alongside increases in mid-range quintiles (40th + 60th from 41.7% to 68.8%), rather than following a monotonic trend (**Table 4**).

**Table 4.** Score-Group Distribution of Returning ENADE Participants by Exam Year.

| Score Groups | 2016<br>(n=161) | 2019<br>(n=175) | 2023<br>(n=221) | Global<br>p-value | Trend<br>p-value | 2016 vs 2019<br>p-value | 2019 vs 2023<br>p-value |
| --- | --- | --- | --- | --- | --- | --- | --- |
| **ENADE** |  |  |  | **<0.001** | **<0.001** | **0.002** | 0.161 |
| 20th | 7 (4.3 %) | 11 (6.3 %) | 6 (2.7 %) |  |  |  |  |
| 40th | 27 (16.8 %) | 17 (9.7 %) | 27 (12.2 %) |  |  |  |  |
| 60th | 70 (43.5 %) | 62 (35.4 %) | 73 (33.0 %) |  |  |  |  |
| 80th | 55 (34.2 %) | 66 (37.7 %) | 77 (34.8 %) |  |  |  |  |
| 100th | 2 (1.2 %) | 19 (10.9 %) | 38 (17.2 %) |  |  |  |  |
| **IDD** |  |  |  | **<0.001** | 0.220 | **<0.001** | **<0.001** |
| 20th | 8 (5.0 %) | 7 (4.0 %) | 12 (5.4 %) |  |  |  |  |
| 40th | 26 (16.1 %) | 13 (7.4 %) | 59 (26.7 %) |  |  |  |  |
| 60th | 79 (49.1 %) | 60 (34.3 %) | 93 (42.1 %) |  |  |  |  |
| 80th | 41 (25.5 %) | 87 (49.7 %) | 51 (23.1 %) |  |  |  |  |
| 100th | 7 (4.3 %) | 8 (4.6 %) | 6 (2.7 %) |  |  |  |  |

**Table Note:** ENADE: National Student Performance Exam; IDD: Indicator of Difference between Observed and Expected Academic Performance.

These results suggest that the pronounced shift toward lower performance in 2023 was driven primarily by first-time participants, possibly reflecting initial resource constraints or unfamiliarity with the examination, while returning institutions maintained a more stable distribution.

### 3.5 Class Size Correlates Negatively with Performance

Larger class sizes possessed a weak negative correlation with lower performance on both ENADE (rho = -0.202, p-value < 0.001) and IDD scores (rho = -0.09, p-value = 0.005), as shown in **Supplementary** Figures 1 and 2. While some subgroup-value analyses showed non-significant trends, the strongest and only statistically significant correlation at the subgroup-value level was observed among public institutions for the IDD score (rho = -0.141, p-value = 0.008). Among for-profit schools, negative trends were observed in both ENADE and IDD, but these did not reach statistical significance. These findings suggest that increasing class sizes may be modestly associated with diminished academic performance, particularly in contexts where institutional resources may be stretched.

### 3.6 Bayesian Regression Analysis: Administrative Category Matters

Finally, Bayesian mixed-effects models, adjusted for both spatial and temporal random effects, further reinforced our earlier findings. Public institutions outscored for-profit universities by an average of 21.5 (95% CrI: 18.6-24.4) ENADE points higher, with non-profit institutions scoring 5.9 points higher (95% CrI: 2.92-8.79) than for-profits. Conversely, for IDD administrative category showed no significant impacts. Instead, class size emerged as a significant negative, albeit modest, predictor (-0.04, 95% CrI: -0.06 to –0.01) (**Table 5**). This finding suggests that, as class sizes increase, the university’s educational value (IDD) tends to decrease at a ratio of -0.04 (-0.06 to -0.01) per student (**Table 5**).

**Table 5.** Bayesian Mixed-Effects Regression Results for ENADE and IDD Scores.

| Variables | ENADE Score |  | IDD Score |  |
| --- | --- | --- | --- | --- |
|  | Regression Coefficients [95% CrI] | Significance | Regression Coefficients [95% CrI] | Significance |
| Private (non-lucrative) | 5.85 [2.91 - 8.79] | Significant | 0.77 [-2.62 - 4.17] | Not Significant |
| Public | 21.52 [18.65 - 24.38] | Significant | 0.84 [-2.31 - 4.22] | Not Significant |
| Class Size | -0.01 [-0.03 - 0.01] | Not Significant | -0.04 [-0.06 - -0.01] | Significant |

**Table Note:** This table displays results from Bayesian mixed-effects regression models fitted using the Integrated Nested Laplace Approximation (INLA) framework, incorporating random effects to account for both state-level and year-specific variations in score values. These two models analysed whether institutional types, as well as class size were associated with ENADE and IDD performance in this controlled environment. For-profit private schools served as the reference group for the institution category, and class size was included as a continuous variable. Regression coefficients are presented with their 95% credible intervals (CrI). A coefficient is deemed statistically significant if its CrI does not include zero, reflecting consistent Bayesian inference within the model.

## 4 Discussion

The findings of this study reveal a concerning association between the accelerated expansion of medical education in Brazil, particularly in the for-profit private sector, and the stagnation or regression of academic performance indicators. In the last decade, undergraduate medical education has grown rapidly in Brazil, especially in the for-profit private sector (2). This expansion led to a sharp rise in student enrollment, particularly in newly established institutions. However, this increase in enrollment did not translate into better academic outcomes. Although the number of ENADE participants increased substantially between 2013 and 2023, average scores remained stagnant, with consistently lower performance among for-profit institutions. Even more concerning was the sharp decline in IDD scores between 2019 and 2023, even after controlling for socioeconomic factors, suggesting a deterioration in the formative capacity of these institutions. This trend points to structural weaknesses, especially in institutions that expanded rapidly without sufficient academic investment.

This rapid expansion was largely shaped by governmental initiatives such as the *Programa Mais Médicos* which aimed to reduce physician shortages in underserved regions by encouraging the creation of new medical schools. However, it also enabled the rapid growth of for-profit institutions, often led by large educational conglomerates (2). However, this expansion was not consistently accompanied by proportional investments in academic quality and infrastructure, resulting in measurable harm to the training of future physicians (1).

Based on the available data, the evidence is unequivocal: institutional growth, particularly in for-profit environments, has outpaced quality assurance mechanisms. It is hypothesized that mass enrollment without multidimensional investments in faculty capacity and research experience has weakened educational outcomes. The rush to scale up medical education has likely left thousands of students underprepared.

This observation reinforces longstanding concerns about the commodification of medical education, in which expansion is driven more by economic interests than by academic rigor or societal needs (1). The proliferation of medical schools, when not accompanied by institutional and pedagogical reform, risks producing graduates ill-equipped to function in complex health systems centered on population needs (16). This deregulated expansion model often results in fragmented curricula, overly hospital-centric training, and insufficient preparation for teamwork, factors that collectively weaken graduates’ readiness to meet modern public health challenges (1,14).

As discussed in our previous work, the entrance of publicly traded educational conglomerates has transformed medicine into a commodity (1). The logic of maximizing profit per student often comes at the expense of mentorship, clinical training, and academic rigor. This dynamic further widens the gap between educational practice and the ethical mission of the profession, undermining both the professional identity and social responsibility of future doctors (16).

Other studies point to similar trends in countries such as India, Peru, and South Africa, where the rapid expansion of private medical schools, often driven by commercial interests, has raised concerns about educational quality (5,6). In the African context, for instance, initiatives such as the Medical Education Partnership Initiative emerged precisely to address challenges stemming from this expansion, including deficiencies in the teaching of basic sciences such as anatomy, physiology, and biochemistry, often exacerbated by a shortage of qualified faculty and limited infrastructure (17).

These deficiencies are compounded by inadequate infrastructure, lack of qualified faculty, and weak integration between foundational sciences and clinical training, which directly undermines the quality of medical education (1,17–19). Such global parallels underscore the urgent need for effective regulatory policies to safeguard quality training amid accelerated institutional growth.

The 2023 evaluation cycle revealed significant disparities across institutional types, with public universities outperforming both non-profit and for-profit private institutions on ENADE scores. Even within the private sector, non-profit institutions tended to outperform for-profit ones, reinforcing the association between institutional governance and the quality of medical education (4,20,21). While IDD scores did not differ significantly between groups, the substantially lower absolute ENADE scores observed in for-profit institutions suggest that any learning gains achieved during training may be insufficient to overcome existing structural limitations.

The IDD metrics were originally designed to estimate the value added by each program, accounting for incoming student background and focusing on institutional impact. However, as previously noted (22), this metric may obscure structural failures in schools that admit high volumes of students but deliver limited pedagogical returns. Given that the recent expansion of medical schools is largely driven by the private sector, and that ENADE is taken at the end of undergraduate training, the 2023 data likely reflects the performance of these new schools (22).

Bayesian mixed-effects regression models strengthened these conclusions by isolating the effects of institutional type and class size. While public institutions demonstrated significantly higher ENADE scores, IDD was not significantly associated with administrative category, highlighting potential limitations in this metric’s sensitivity to institutional quality. In contrast, class size was a significant negative predictor of IDD scores, underscoring the importance of linking enrollment growth to proportional increases in infrastructure and faculty support. Further analysis of class-size-normalized scores reinforced these findings. Even when controlling for cohort size, public institutions maintained superior performance. This suggests that institutional type matters not only in terms of scale, but also due to underlying structural and pedagogical differences.

This model becomes particularly urgent when considering basic science education, whose historical fragmentation and theoretical excess have drawn criticism from recent curricular reform movements As previously discussed (1), the erosion of basic science training, characterized by cuts in course hours, reduced access to experimental projects, and the devaluation of non-physician faculty, undermines scientific literacy and clinical reasoning, both essential to safe and effective medical practice.

The sharp decline in performance observed among first-time ENADE participants in 2023 reflects a recurring pattern of prematurely launched medical schools lacking institutional maturity. These findings emphasize the need for more rigorous regulatory criteria for opening new programs, including minimum standards for infrastructure, qualified faculty, and coherent pedagogical planning.

By compromising training quality, especially in underserved or rural regions, unregulated expansion undermines the very goals of initiatives like the *Programa Mais Médicos*, which aimed to increase access to medical services for vulnerable populations. The findings of this study underscore the urgent need to reassess public policies governing medical education in Brazil. While initiatives such as the *Programa Mais Médicos* sought to increase access to medical degrees, the evidence presented here suggests that unregulated expansion without stringent quality controls may result in substandard training, with potentially severe consequences for the healthcare system.

Medical education in the 21st century must be guided by principles of transformative learning and institutional interdependence, rejecting the fragmented, commodified model still prevalent in many countries (1,16). Future policies must combine expansion mechanisms with robust regulation, rigorous accreditation, transparency in performance metrics, and a steadfast commitment to the social mission of medical education. Training future physicians must serve the public interest, promote equity in access to care, and ensure the excellence needed to prepare committed professionals.

The expansion, privatization of medical education in Brazil demand more rigorous and consistent monitoring. The findings presented here highlight the critical need to not only train more physicians but also ensure that they receive high-quality education. Only then will it be possible to effectively improve healthcare access for the Brazilian population.

To translate these findings into effective action, we suggest policy measures designed to safeguard and ultimately improve the quality of medical training in Brazil. The evidence presented above indicates how unregulated growth, especially in for-profit programs, has outpaced investments in faculty, infrastructure, and curricular coherence, leading to stagnation in ENADE outcomes and decline in the educational value (IDD) of medical schools. Considering these insights, we offer the policy recommendations (**Box 1**), each grounded in our empirical analysis, to realign expansion with educational excellence and the public health commitment of medical education in Brazil.

Recent developments underscore the urgency of the findings reported here. In April 2025, the Ministry of Education launched the National Examination for the Evaluation of Medical Education (ENAMED) (23). Inspired by the National Residency Exam (ENARE), the exam will be administered annually to graduating students. According to the Ministry of Education (MEC) and Ministry of Health (MoH), The main goal of the newly established ENAMED is to shift the responsibility for poor student performance to institutions, especially private for-profit programs, which have concentrated the worst ENADE scores. In 2023, 27.3% of private medical courses received grades 1 or 2, compared to just 6% of public institutions, prompting the declaration from the MEC: “students are not responsible for their training, institutions are.” (24).

We believe that ENAMED signals a new phase in Brazil’s medical education policy. Oversight and performance monitoring are expected to increase, particularly in institutions that have grown rapidly without ensuring quality. Whether the initiative will reverse current trends remains to be seen, but it clearly aligns with the need for systemic accountability identified in this analysis.

This study has some limitations. Although this study relies on comprehensive and robust national datasets, it is limited by the absence of qualitative variables that could provide insight into institutional culture, teaching practices, and student support. Future research should incorporate qualitative methods and longitudinal tracking of graduates to better understand how institutional differences shape professional development.

## 5 Conclusion

Our comprehensive analysis of nationwide educational data reveals a troubling decline in the academic performance of medical graduates from private institutions in Brazil—most notably within the for-profit sector. The deeper crisis in private medical education is not limited to its commercialization; it lies in the steady dismantling of medicine’s ethical and social foundations.

When expansion prioritizes volume over rigor, profit over purpose, and growth over equity, the consequences reach far beyond the classroom—placing both students and future patients at risk. This is more than a policy misstep; it is a profound breach of public trust. To safeguard the future of healthcare in Brazil, medical education must be reoriented toward excellence, integrity, and a genuine commitment to public service. The sustainability of our health system depends on our collective capacity to realign training with its core mission—so that the expansion of medical schools strengthens, rather than weakens, the nation’s commitment to health and human dignity.

## 7 Author Contributions

BBA: Conceptualization, Supervision, Writing - original draft and Writing - review and editing. KVS: Formal Analysis, Writing - original draft and Writing - Review and Editing. RCM: Formal Analysis, Writing - original draft and Writing - Review and Editing. LFQ: Writing - original draft and Writing - Review and Editing. KMA: Writing - original draft and Writing - Review and Editing.

## Data Availability

All data produced in the present study are available upon reasonable request to the authors

## Acknowledgments

This is a short text to acknowledge the contributions of specific colleagues, institutions, or agencies that aided the efforts of the authors.

## 9 Data Availability Statement

The datasets analyzed for this study are publicly available at: https://www.gov.br/inep/pt-br/acesso-a-informacao/dados-abertos/indicadores-educacionais/indicadores-de-qualidade-da-educacao-superior

## Box 1. Policy Recommendations

1. Implement Longitudinal Evaluations: Periodic formative assessments should monitor learning throughout medical training.
2. Reform and Strengthen Accreditation Criteria: Institutions must be evaluated based on faculty/student ratios, infrastructure, and historical performance.
3. Align Expansion with Health Needs: Incentivize regional equity without compromising on quality.
4. Adopt a National Licensing Exam: To ensure minimal competency standards for medical graduates.

