## Supplemental Material for "For-Profit Growth and Academic Decline: A Retrospective Nationwide Assessment of Brazilian Medical Schools"

### *Supplementary Material*

#### 1 Supplementary Figures

**Supplementary Figure 1.** Correlation Between ENADE Scores and Class Sizes.

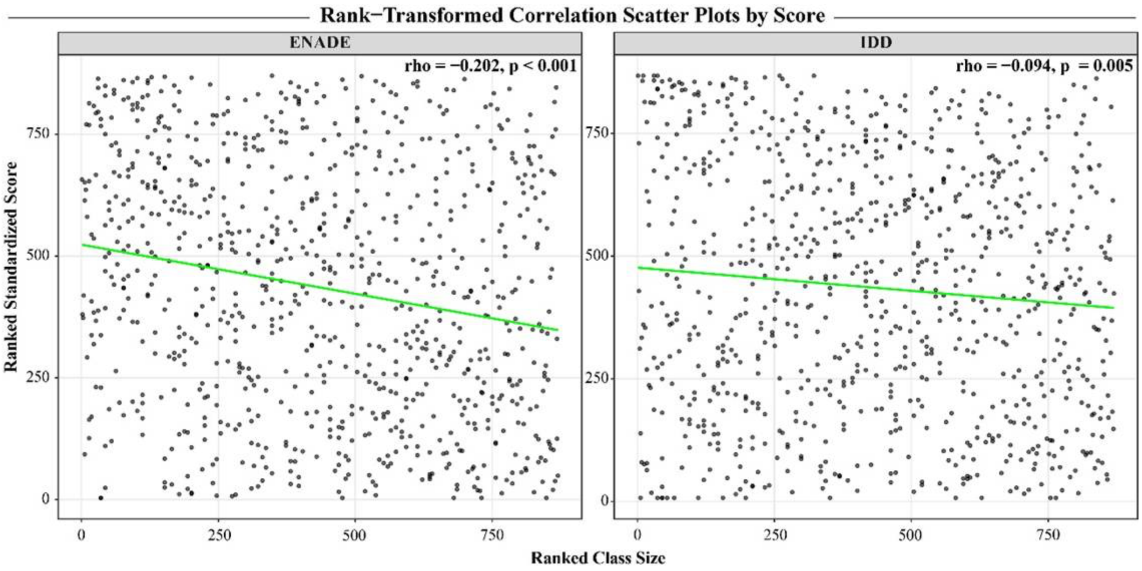


**Figure Legend:** Spearman Correlation scatter plots assessing the relationship between medical school class sizes and standardized scores for continuous ENADE and IDD scores. Ranked-transformed data on both standardized scores, as well as class sizes, is presented with the green line representing the direction of association. Spearman's rho and respective p-values are presented in text.

**Supplementary Figure 2.** Correlation Between ENADE Scores and Class Sizes According to Institutional Types.

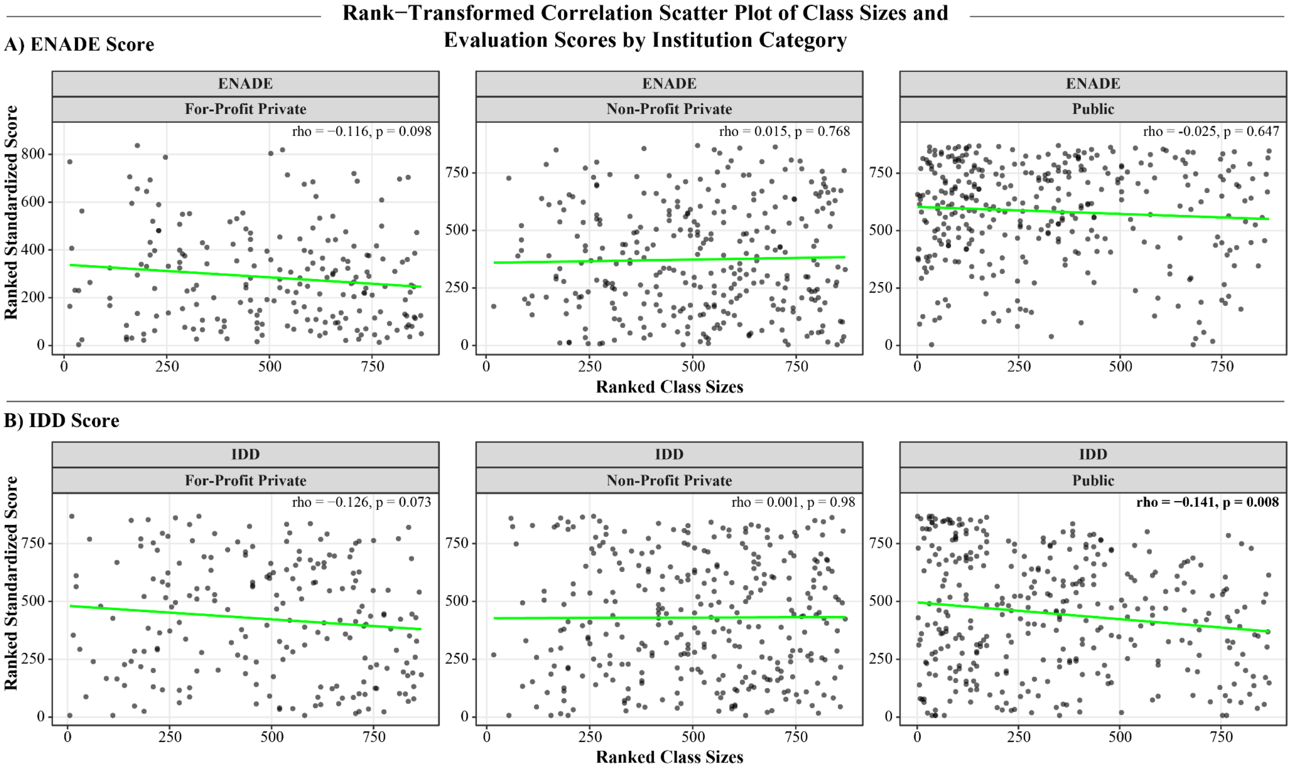


**Figure Legend:** Spearman Correlation scatter plots assessing the relationship between medical school class sizes and university standardized scores for continuous ENADE and IDD scores, according to their institutional types (for-profit private, non-profit private, and public institutions). Ranked-transformed data on both standardized scores, as well as class sizes, is presented with the green line representing the direction of association. Spearman's rho and respective p-values are presented in text.
